# The correlation between lumbar and pelvic dysfunctions in non-post-surgical adult patients: a scoping review protocol

**DOI:** 10.1101/2024.05.02.24305675

**Authors:** Tiziana Manni, Francesca Coaro, Monica Nativo, Marina Stocco

## Abstract

**Objective:** The objective of this scoping review is to understand the extent and type of evidence available on the correlation between pelvic and lumbar region dysfunction in adult non-surgical patients.

**Introduction:** It is well known that the pelvic and lumbar regions are connected, but the information about them is conflicting and often not useful for clinical practice. For this reason, we want to conduct a scoping review, to map all available evidence on the topic and provide clearer information for clinical work with on patients.

**Inclusion criteria:** This scoping review will consider adult patients with pelvic floor and and low back disfunction with related clinical conditions. The only limitation is post-surgical patients, who will be excluded from this review. This scoping review will consider any study design or type of publication.

**Methods:** An initial limited search of MEDLINE (PubMed) will be undertaken followed by analysis of the text words contained in the title and abstract, and of the index terms used to describe these articles. This will lead to the development of a search strategy including identified keywords and index terms that fit each included database: CINAHL, Cochrane Library, PEDro, Scopus and grey literature. The research will be performed from inception to February 2024. No geographical, setting and language restrictions will be applied. Potentially relevant sources will be retrieved in full, and their citation details imported into Rayyan — a web and mobile app for systematic reviews. Additional records will be identified through searching in grey literature and the reference lists of all relevant studies. No study design, publication type, data and language restrictions will be applied. Two reviewers will independently screen all abstracts and full-text studies for inclusion. A data collection form will be used to extract the characteristics of the studies included. A tabular and accompanying narrative summary of the information will be provided. The results of the search and the study inclusion process will be presented in a Preferred Reporting Items for Systematic Reviews and Meta-analyses extension for scoping review (PRISMA-ScR) flow diagram.

## Introduction

A preliminary search of MEDLINE, the Cochrane Database of Systematic Reviews and *JBI Evidence Synthesis* was conducted and no current or underway scoping reviews on the topic were identified. Some systematic reviews exist, but they compare specific intervention treatments, which do not allow to have a global overview of the subject. Therefore, the need to collect comprehensively all the available evidence on the correlation between lumbar and pelvic dysfunctions was highlighted. For this reason, this scoping review aims to include all the useful information available in literature including all assessment and treatment methods, all assessment tools, all treatment and all outcome measures. The population considered will be of non-post-surgical adult patients with no limits of settings, time, geographical regions, ethnicity and sex.

The pelvic floor consists of fascias, ligaments and the pelvic floor muscles (PFM). The PFM is composed of twelve muscles organized into three layers that extend from the pubic symphysis to the walls of the ilium and coccyx [1]. All these muscles form the basis of the abdominal cavity on which the pelvic organs rest and where the urethra, vagina and rectum pass; they therefore guarantee urinary and fecal continence, support the pelvic viscera and contribute to the control of intra-abdominal pressure [2]. There are also some evidence about their role in the stabilization of the sacro-iliac joint [3] but the evidence on this is conflicting [4]. For this reason, a review that clearly defines what knowledge on the subject is demonstrated in the literature is necessary.

## Review question

To provide a comprehensive picture of lumbo-pelvic dysfunction, this scoping review aims to identify and present the available information regarding the correlation that exists between pelvic and lumbar region dysfunction; this in order also to identify knowledge gaps in this field.

Therefore, the main question of this review is: what evidence is available about the correlation between lumbar and pelvic dysfunctions in non-post-surgical adult patients?

## Eligibility criteria

Studies will be eligible for inclusion if they meet the following Population, Concept and Context (PPC) criteria.

### Population

This scoping review will take into consideration adults patients with pelvic floor and and low back disfunction with related clinical conditions. No surgical conditions will be included. This exclusion is made to have a focus only on patients treated conservatively, which potentially have very different characteristics from surgical ones. Instead, pregnant women will be included in the research.

### Concept

Outcome measures, assessment tools, treatments and results examined by the researchers will be considered.

### Context

This review will consider studies conducted in any context.

### Types of Sources

This scoping review will consider any study design or publication type. No time, geographical, setting and language restrictions will be applied.”

## Methods

The proposed scoping review will be conducted in accordance with the JBI methodology for scoping reviews: the Preferred Reporting Items for Systematic reviews and Meta-Analyses extension for Scoping Reviews (PRISMA-ScR) [6].

### Search strategy

The search strategy will aim to locate both published and unpublished studies. An initial limited search of MEDLINE (PubMed) was undertaken to identify articles on the topic. The text words contained in the titles and abstracts of relevant articles, and the index terms used to describe the articles were used to develop a full search strategy for CINAHL, Cochrane Library, PEDro, Scopus. (see Appendix I). The search strategy, including all identified keywords and index terms, will be adapted for each included database and/or information source.

Studies published in any language will be included. In case of articles in languages different from Italian, English or Spanish, will be the authors’ responsibility to hire a translator. No publication date limitations will be applied.

Also, sources of unpublished studies and grey literature will be search.

### Source of Evidence selection

Following the search, all identified citations will be collated and uploaded into EndNote 21.2/2023 (Clarivate Analytics, PA, USA) and duplicates removed. Following a pilot test, titles and abstracts will then be screened by two independent reviewers for assessment against the inclusion criteria for the review. Potentially relevant sources will be retrieved in full and their citation details imported into the JBI System for the Unified Management, Assessment and Review of Information (JBI SUMARI) (JBI, Adelaide, Australia) [5]. The full text of selected citations will be assessed in detail against the inclusion criteria by two other independent reviewers. Reasons for exclusion of sources of evidence at full text that do not meet the inclusion criteria will be recorded and reported in the scoping review. Any disagreements that arise between the reviewers at each stage of the selection process will be resolved through discussion, or with an additional reviewer/s.

### Data Extraction

Data will be extracted from papers included in the scoping review by two independent reviewers using the JBI data extraction tool. The data extracted will include specific details about the participants, concept, context, study methods and key findings relevant to the review question. A draft extraction form is provided (see Appendix II). It will be modified and revised during the process of extracting data from each included evidence source, if necessary. Modifications will be detailed in the scoping review. Any disagreements that arise between the reviewers will be resolved through discussion, or with two additional reviewers.

If appropriate, authors of papers will be contacted to request missing or additional data, where required.

### Data Analysis and Presentation

The evidence presented will directly respond to the review objective and question. The data will be presented mostly in tabular form and a narrative summary will accompany the tabulated to describe how the results relate to the reviews objective and question.

This review will present the available information regarding the correlation between the dysfunctions of the lumbar region and the ones of the pelvic region; the findings from this review will be useful to the clinical practice of physiotherapists who deal with patients with the dysfunctions presented in this review.

## Data Availability

All data produced in the present study are available upon reasonable request to the authors

## Acknowledgements

Francesca Coaro – Università degli Studi di Padova

## Appendix I: Search strategy

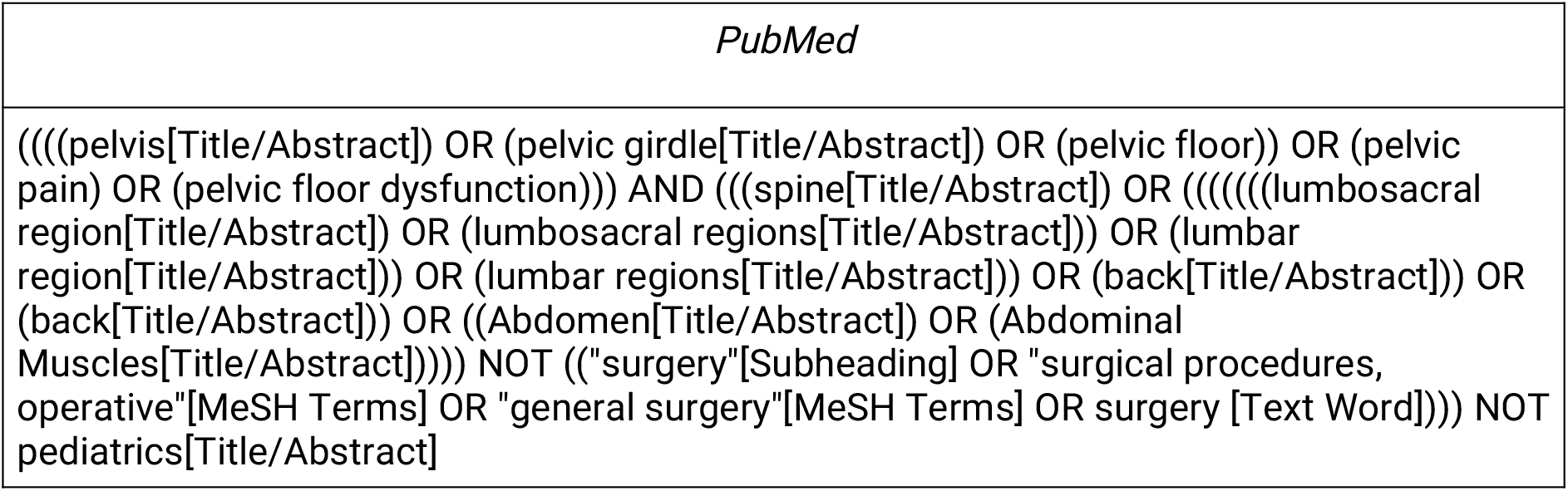

## Appendix II: Data extraction tool

|  |
| --- |
| Author(s) and year of publication |
| Origin/country of origin (where the source was published or conducted) |
| Aims/purpose |
| Population/sample size (if applicable) |
| Methodology/methods |
| Intervention type and duration, comparater |
| Outcome measures |
| Key findings that relate to the scoping review question/s |

